# Social Media Platforms for Health Communication and Research in the Face of COVID-19 Pandemic: A Cross Sectional Survey in Uganda

**DOI:** 10.1101/2020.04.30.20086553

**Authors:** Ronald Olum, Felix Bongomin

## Abstract

**Objectives:** (1) To examine the usage of social media and other forms of media among medical students (MS) and healthcare professionals (HCPs) in Uganda. (2) To assess the perceived usefulness of social media and other forms of media for COVID-19 public health campaigns.

**Design:** A descriptive WhatsApp messenger-based cross-sectional survey in April 2020.

**Setting:** Makerere University Teaching Hospitals (MUTH) and 9 of the 10 medical schools in Uganda.

**Participants:** HCPs at MUTH and MS in the 9 medical schools in Uganda.

**Main outcome measures:** We collected data on sociodemographic characteristics, sources of information on COVID-19, preferences of social media platform and perceived usefulness of the different media platforms for acquisition of knowledge on COVID-19.

**Result:** Overall, response rate was 21.5% for both MS and HCPs. In total, 877 (HCPS [136, 15.5%], MS [741, 85.5%]) were studied. Majority (n=555, 63.3%) were male with a median age of 24 (range: 18-66) years. Social media was a source of information for 665 (75.8%) participants. Usage was similar among MS and HCPs (565/741 (76.2%) vs. 100/136 (73.5%), p=0.5). Among the MS, commonly used social media were: WhatsApp (n=705, 95.1%) Facebook (n=405, 54.8%), Twitter (n=290, 39.1%), Instagram (n=178, 24.0) and Telegram (n=80, 10.8%). Except for WhatsApp, male MS we more likely to use the other social media platforms (p= <0.001 − 0.01). Mass media (television and radio) and social media were preferred the most useful tools for dissemination of COVID-19 related information.

**Conclusion:** More than two-thirds of MS and HCPs are routinely using social media in Uganda. Social media platforms may be used for dissemination of information as well as a research tool among MS and HCPs. Social media alongside other media platforms can also be used as sources of reliable information on COVID-19 as well as for dissemination of research findings and guidelines.

Strengths and limitations
- This is the first study in sub Saharan Africa on the use of social media for research during the COVID-19 pandemic.
- The study also explores perceived usefulness of different media for COVID-19 public health campaigns.
- Diversity of the participants consisting both healthcare professionals and medical students.
- A relatively large sample size was enrolled in the survey despite a low response rate.

## INTRODUCTION

First reported in Wuhan, China in December and declared a pandemic by World Health Organisation (WHO) on March 21^st^ 2020, coronavirus disease 2019 (COVID-19) caused by a novel human coronavirus (SARS-CoV-2) has rapidly spread to over 200 countries and territories globally [1–3]. The on-going COVID-19 pandemic has threatened the lives of over 3 million people, claiming over 200,000 lives worldwide [3, 4].

Researchers globally are racing to identify an effective vaccine and treatment for the viral disease, in order to curb the high morbidity and mortality associated with this virus. WHO has recommended maintaining a social distance universally to reduce human to human transmission of COVID-19 [5]. As a result, there has been widespread lockdown in most countries in a bid to reduce public gatherings and rapid spread of the disease [6]. This has affected nearly all sectors, the health sector not spared. Except for COVID-19 related studies, other biomedical researches that involve contacts with participants onsite have reduced significantly in many countries [7, 8]. Researchers have been advised to utilise virtual means including teleconferencing, virtual lab meetings and research seminars to maintain studies that can be conducted remotely [7].

Health campaigns aimed at increasing the awareness of the public on transmission and prevention of the virus are also being employed by various international and local organisations. Mass media and social media have been frequently used to disseminate infographics on the pandemic[9].

In this study, we explored the usage and perceived usefulness of social media and other forms of media among medical students (MS) and healthcare professionals (HCPs) in Uganda.

## METHODS AND MATERIALS

### Study design

We conducted an online, descriptive cross-sectional study between Wednesday 1^st^ April and Sunday 19th April 2020 as part of a larger study assessing knowledge, attitude and practices towards COVID among healthcare workers [10] and medical students. A quantitative analysis approached was used.

### Study settings

Participants were derived from 2 settings: 1) Medical students from 9 of the 10 universities in Uganda offering undergraduate medical degrees with a combined population of about 6,000 students. 2) Health care professionals from Makerere University Teaching Hospitals (MUTHs), with a population size of about 1,300-1,500.

### Study Population

Medical students pursuing Bachelor of Medicine and Bachelor of Surgery, Bachelor of Dental Surgery, Bachelor of Nursing and Bachelor of Pharmacy and Healthcare professionals including nurses, midwives, intern doctors, medical officers, residents and specialists at the MUTH. Individuals aged 18 years or older were included in the study after an informed consent was obtained. Students and healthcare professionals who were too ill or were offline during the time of the study were excluded.

### Study sample and procedure

By employing convenience-sampling method, we used WhatsApp Messenger (Facebook Inc., California, USA) for enrolling potential participants. We identified all the existing WhatsApp groups of medical students in the various universities and those of healthcare professionals in the different MUTHs. A total of about 3,500 students and 581 healthcare professionals who were members in the several WhatsApp groups were approached to participate in the study. An online data collection tool was designed and executed using Google Forms (via docs.google.com/forms). The Google Form link to the questionnaire was sent to the enrolled participants via the identified WhatsApp groups.

### Study variables

Independent variables were demographic characteristics were sex, age, and sources of information on COVID-19 and dependent variables were usage and perceived usefulness of social media.

### Data analysis

Microsoft Excel 2016 was used for data cleaning and coding and STATA version 15.1 (StataCorp, College Station, TX) for analyses. Numerical data was analysed using parametric or non-parametric approaches as appropriate. Categorical data was summarized as frequencies and proportions and associations between independent and dependent variables were assessed using chi-square test and logistic regression. A *P< .05* is considered statistically significant.

## RESULTS

Overall, we achieved a response rate of 21.5% (877/4081: 741/3,500 (MS) and 136/581 (HCPs)). Majority of the participants were male (n=555, 63%) and medical students (n=741, 84%). The median age of the participants was 24 (range: 18-66) years

### Source of information

Mass media (n=681, 78%) and social media (n=665, 76%) were the most used sources of information. Female participants were less likely to use journals and websites than male participants. Medical students significantly used mass media like TV (aOR: 1.7, 95% CI: 1.0-2.7, *P=.031*) but were less likely to use websites (adjusted odds ratio (aOR): 0.1, 95% CI: 0.1-0.2, *P<.001*) compared to HCPs to access information on COVID-19, **Table 1**.

**Table 1.** Multivariate analysis showing factors associated with sources of information on COVID-19 among Ugandan medical students and HCPs at Makerere University Teaching Hospitals

| Sources of information (N=877) |  | Freq (%) | aOR (95% CI) | P-value |
| --- | --- | --- | --- | --- |
| Social Media |  | 665 (76) |  |  |
| Sex | Male | 427 (77) | 1.0 |  |
|  | Female | 238 (74) | 0.9 (0.6 - 1.2) | .46 |
| Age | 18 to 23 | 316 (74) | 1.0 |  |
|  | ≥24 | 349 (78) | 1.3 (0.9 - 1.9) | .13 |
| Profession | HCP | 100 (74) | 1.0 |  |
|  | Medical student | 565 (76) | 1.4 (0.8 - 2.2) | .21 |
| Mass Media (TVs) |  | 681 (78) |  |  |
| Sex | Male | 430 (77) | 1 |  |
|  | Female | 251 (78) | 1.1 (0.8 - 1.5) | .69 |
| Age | 18 to 23 | 328 (77) | 1 |  |
|  | ≥24 | 353 (78) | 1.3 (0.9 - 1.9) | .15 |
| Profession | HCP | 98 (72) | 1 |  |
|  | Medical student | 583 (79) | 1.7 (1 - 2.7) | .03 |
| Journals |  | 355 (40) |  |  |
| Sex | Male | 253 (46) | 1 |  |
|  | Female | 102 (32) | 0.6 (0.4 - 0.7) | <.001 |
| Age | 18 to 23 | 163 (38) | 1 |  |
|  | ≥24 | 192 (43) | 1.0 (0.8 - 1.4) | .91 |
| Profession | HCP | 63 (46) | 1 |  |
|  | Medical student | 292 (39) | 0.8 (0.5 - 1.1) | .19 |
| Websites |  | 479 (55) |  |  |
| Sex | Male | 325 (59) | 1 |  |
|  | Female | 154 (48) | 0.6 (0.5 - 0.9) | .003 |
| Age | 18 to 23 | 194 (45) | 1 |  |
|  | ≥24 | 285 (63) | 1.2 (0.9 - 1.6) | .288 |
| Profession | HCP | 125 (92) | 1 |  |
|  | Medical student | 354 (48) | 0.1 (0.0 - 0.2) | <.001 |

### Preferred social media usage among medical students

About 95% of medical students frequently used WhatsApp followed by Facebook (55%) and only 1% (n=4) did not frequently use any social media platform, ***Figure 1***. Excluding WhatsApp, male students were more likely to use the other social media platforms than female medical students (*P-values* between <.001 and 0.01). Medical students ≥24 years were more likely to use Facebook (aOR: 1.8, 95% CI: 1.4 to 2.5, *P<.001*) but less likely to use Instagram (aOR: 1.7, 95% CI: 1.0-2.7, *P=.031*), ***Table 2***.

**Figure 1.**
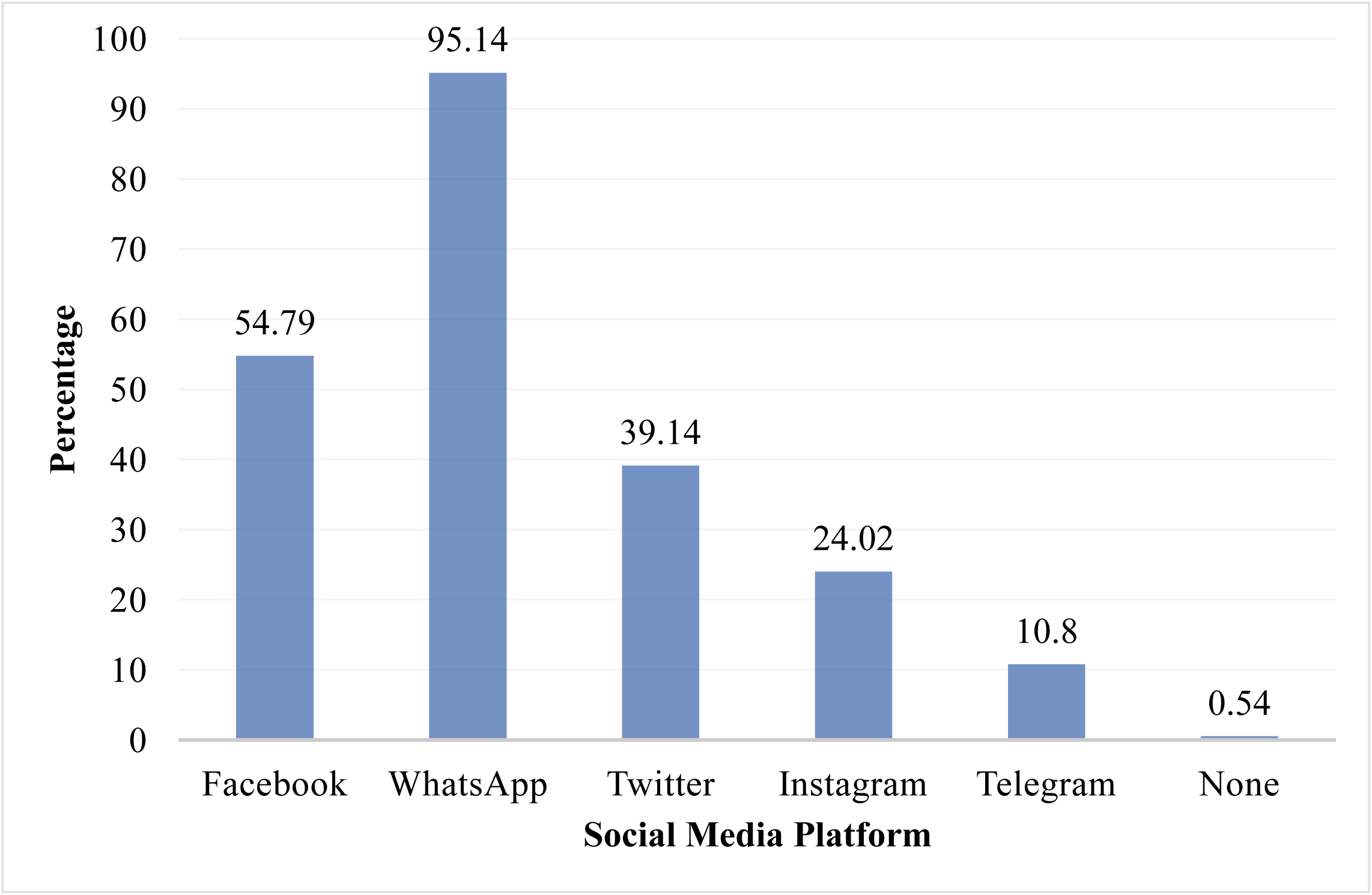
Frequently used social media platforms among medical students in Uganda

**Table 2.**
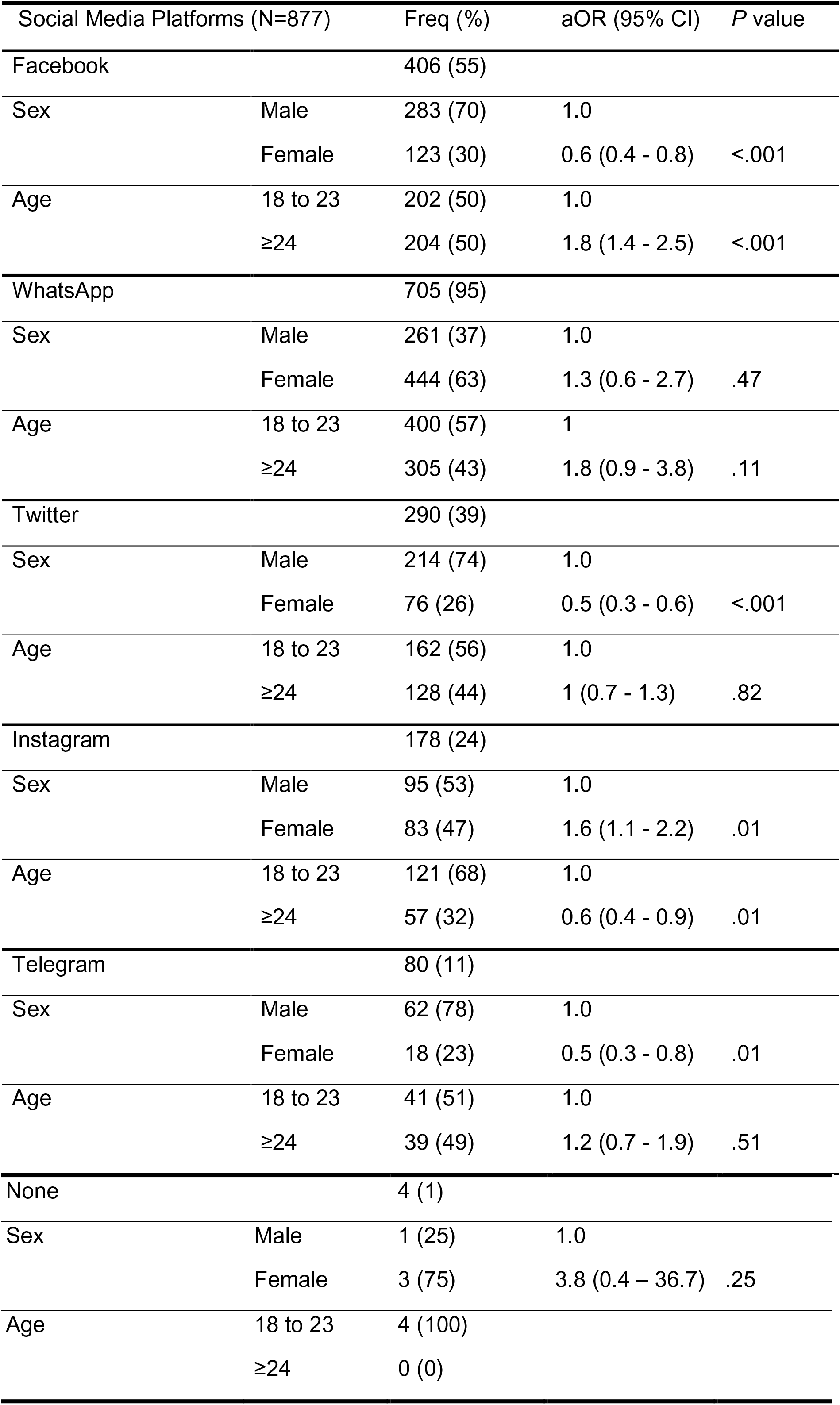
Multivariate analysis showing factors associated with social media use among Ugandan medical students.

### Perceived usefulness of different media for COVID-19 health campaigns

Majority of the medical students recognised television, radios and social media as the most useful tools for dissemination of information of COVID-19, ***Figure 2***. Print media (billboards, banners and newspapers) was perceived as the least useful in COVID-19 public health campaigns.

**Figure 2.**
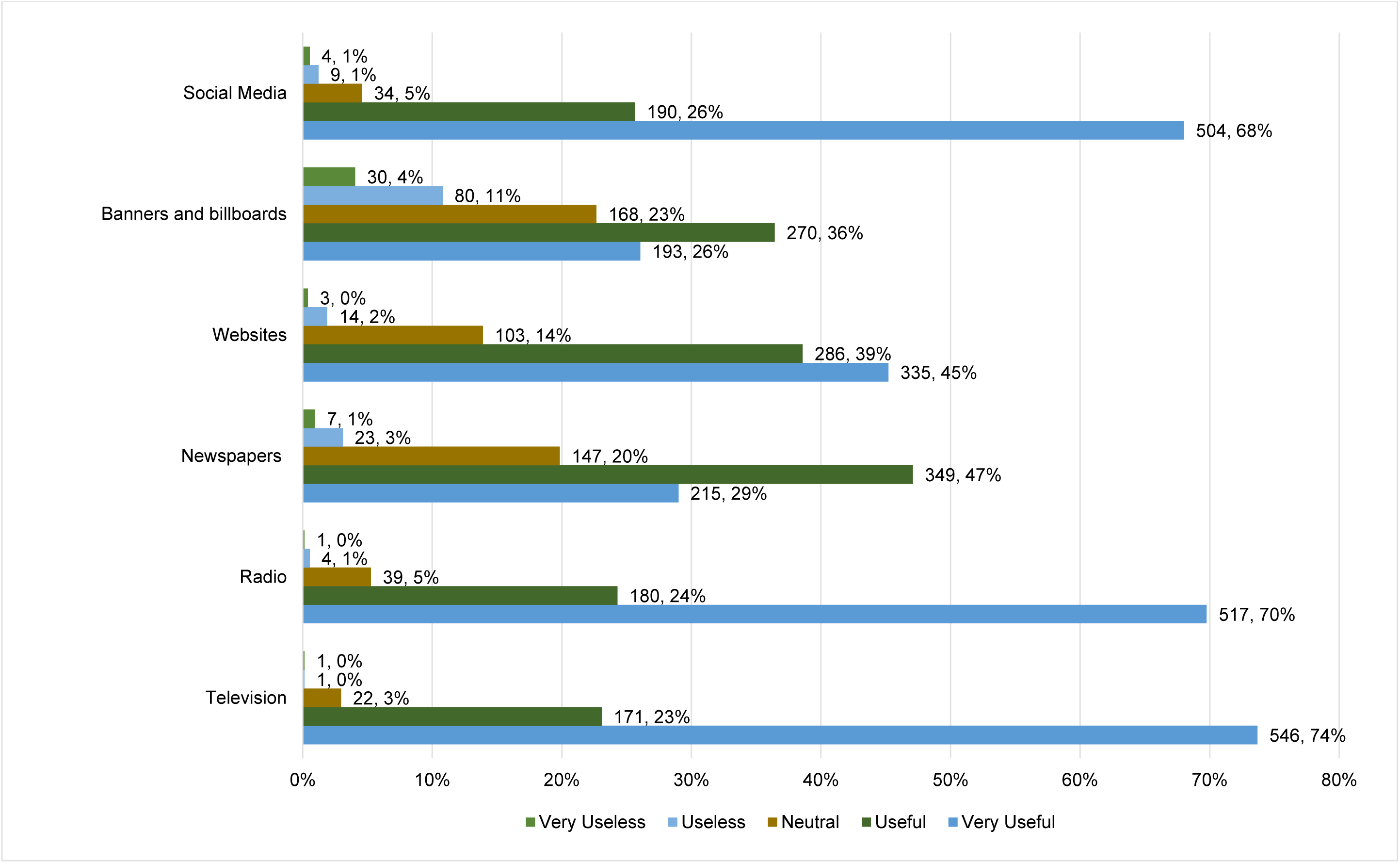
Perceived usefulness of different media for COVID-19 health campaigns by Ugandan medical students

## DISCUSSION

The purpose of the study was to assess the usage and perceived usefulness of social media and other forms of media among medical students and healthcare professionals during the COVID-19 pandemic in Uganda. The results suggest that mass media and social media were the most used sources of information for COVID-19 among HCPs and MS. Among medical students, WhatsApp and Facebook were the most frequently used social media platforms; and they preferred mass media and social media as the most useful tools for dissemination of COVID-19 related information. This is the first study in sub-Saharan Africa assessing social media usage and perceived usefulness of various media for health campaigns during COVID-19 pandemic.

### Public health campaigns

COVID-19 pandemic is currently the greatest public health concern affecting majority of countries globally. Social distancing guidelines and lockdowns have also posed a challenge to public health campaigns. This therefore necessitates a shift from popular print media (newspapers, magazines, banners, etc.) to wireless media. Our study suggests that common wireless media like televisions, radios and social media can be effective in improving awareness on COVID-19. Over 40% of the world’s population have access to internet to date [11]. Social media are the most used networking sites globally, with Facebook being the most used social networking platform [12]. Social media can be used for dissemination of knowledge and clearing myths the public has on COVID [9]. However, misinformation can be equally spread by social media leading to fear, panic and anxiety among the public[13]. Infographics can be therefore be widely distributed via these platforms by verified pages and accounts of public agencies and health officials. WHO already has dedicated WhatsApp numbers and groups in various languages to disseminate info on COVID-19 [14].

Social media can also be used for reporting probable cases, tracing of contacts, making appointments for tests and delivery of test results to the tested clients. Ministry of Health Uganda has dedicated contacts for the public to report any suspected case of COVID-19 to the officials who follow up and perform tests when required [15]. These contacts can also be accessed through WhatsApp.

### Social media and research in COVID-19 times

Through WhatsApp we were able to reach out to over 4,000 medical students and health care professionals within 2 weeks. Despite physical social distancing, the society and the world at large are now largely communicating through social media. We were able to achieve a response rate of 21.5% which is in line with a meta-analysis that reported a range of 7% to 88% [16]. It is however lower than the average response rate in the above study (34%) and among physicians (35%) by Cunningham and colleagues [16, 17]. A few studies conducted using online surveys during the COVID-19 pandemic have also reported promising responses [18, 19]

In addition to the COVID-19 pandemic, Sub-Saharan Africa has a high burden of other ongoing pandemics, notably HIV/AIDS and TB [20]. A vast majority of these patients are on regular follow-ups for routine clinical care and for research purpose. Clinicians and researchers can utilise the widely available social media platforms to conduct interviews and follow-ups of these patients. Chronic care patients who require uninterrupted supply of medications during this lockdown can also maintain communications with their clinical care and research teams through social media platforms.

However, ethical concerns that have been discussed before regarding privacy and confidentiality have to be greatly taken into consideration [21]. Privacy policies and guidelines must be developed by the research teams in conjunction with their respective institutional review boards to safeguard transfer of potentially identifying data.

The study has some limitations. The relatively low response rates limit generalizability. Follow up reminders were sent to the prospective participants to improve responses.

### Conclusions

In conclusion, we have been able to show that social media can be robustly used to collect research data among medical students and health care professionals with high response rates. Beyond being a research tool, social media alongside other media platforms can be used as sources of reliable information on COVID-19 as well as for dissemination of research findings and guidelines.

## Data Availability

The dataset presented in this study can be found publicly online on FigShare.

https://doi.org/10.6084/m9.figshare.12226952

## Authors’ contributions

RO and FB conceptualised, designed the protocol, collected and analysed the data, drafted the manuscript, revised and approved the final version.

## Acknowledgement

Ms Sarah Apoto for clerical work. All the medical students and healthcare professionals who participated in this study.

## Ethical approval

The study was cleared by Mulago Hospital Research Ethics Committee protocol number MHREC 1866. All participants provided an online consent to the study.

## Data availability statement

The dataset presented in this study can be found on FigShare at https://doi.org/10.6084/m9.figshare.12226952

